# Allopurinol and Blood Pressure Variability Following Ischemic Stroke and Transient Ischemic Attack: A Secondary Analysis of XILO-FIST

**DOI:** 10.1101/2023.06.29.23292066

**Authors:** Alexander Stuart MacDonald, Michele Robertson, David Alexander Dickie, Phillip Bath, Kirsten Forbes, Terence Quinn, Niall M Broomfield, Krishna Dani, Alex Doney, Keith W Muir, Allan Struthers, Matthew Walters, Mark Barber, Ajay Bhalla, Alan Cameron, Alexander Dyker, Paul Guyler, Ahamad Hassan, Mark Kearney, Breffni Keegan, Lakshmanan Sekaran, Mary Joan Macleod, Marc Randall, Louise Shaw, Ganesh Subramanian, David Werring, Alex McConnachie, Jesse Dawson

**Affiliations:** School of Medicine, Dentistry and Nursing, College of Medical, Veterinary and Life Sciences, University of Glasgow, UK; Robertson Centre for Biostatistics, School of Health and Wellbeing, College of Medical, Veterinary & Life Sciences, University of Glasgow, Glasgow, G12 8QQ, UK; DD Analytics Cubed Ltd, 73 Union Street, Greenock, Scotland, PA16 8BG, UK; School of Cardiovascular and Metabolic Health, College of Medical, Veterinary & Life Sciences, University of Glasgow, Queen Elizabeth University Hospital, Glasgow, G51 4TF, UK; Stroke Trials Unit, Mental Health & Clinical Neuroscience, University of Nottingham, Nottingham NG7 2UH; Department of Neuroradiology, Institute of Neurological Sciences, Queen Elizabeth University Hospital, 1345 Govan Road, Glasgow, G51 4TF; School of Cardiovascular and Metabolic Health, College of Medical, Veterinary & Life Sciences, University of Glasgow, Glasgow Royal Infirmary, Glasgow, UK; Department of Clinical Psychology and Psychological Therapies, Norwich Medical School, University of East Anglia, NR4 7TJ, UK; Department of Neurology, Institute of Neurological Sciences Glasgow, Queen Elizabeth University Hospital, 1345 Govan Road, Glasgow, G51 4TF, UK; Medicine Monitoring Unit (MEMO), School of Medicine, University of Dundee. Ninewells Hospital, Dundee, DD1 9SY; Division of Imaging and Science Technology, School of Medicine, Ninewells Hospital, Dundee DD1 9SY; School of Neuroscience and Psychology, College of Medical, Veterinary & Life Sciences, University of Glasgow, Queen Elizabeth University Hospital, Glasgow, G51 4TF, UK; Division of Molecular and Clinical Medicine, University of Dundee, UK; University Department of Stroke Care, University Hospital Monklands, Airdrie, ML6 OJS; Department of Stroke, Ageing and Health, Guy’s and St Thomas NHS Foundation Trust, St Thomas’ Hospital, Lambeth Palace Rd, London, SE1 7EH, UK; Wolfson Unit of Clinical Pharmacology, Royal Victoria Infirmary, Newcastle Upon Tyne, UK; Department of Stroke Medicine, Mid and South Essex University Hospitals Group, Southend University Hospital, Prittlewell Chase, Westcliff-on-Sea, Essex, SS0 0RY, UK; Department of Neurology, Leeds General Infirmary, Leeds, UK; The University of Leeds, Leeds, UK; Department of Medicine, South West Acute Hospital, Enniskillen. BT74 6DN, UK; Department of Stroke Medicine. The Luton and Dunstable University Hospital, Bedfordshire, NHSFT, Lewsey Road, Luton, LU4 0DZ, UK; Institute of Medical Sciences, University of Aberdeen, Aberdeen, UK; Department of Neurology, Leeds Teaching Hospitals NHS Trust, Leeds, UK; Department of Stroke Medicine, Royal United Hospital, Combe Park, Bath, BA1 3NG, UK; Department of Stroke Medicine, Nottingham University Hospitals, Nottingham, NG5 1PB, UK; Stroke Research Centre, UCL Queen Square Institute of Neurology, London, UK; Comprehensive Stroke Service, National Hospital for Neurology and Neurosurgery, Queen Square, University College Hospitals NHS Foundation Trust, London, UK

## Abstract

**Background:** Blood Pressure Variability (BPV) is associated with cardiovascular risk and serum uric acid level. We investigated whether BPV is lowered by allopurinol and whether it is related to markers of cerebral small vessel disease.

**Methods:** We used data from a randomised, double-blind, placebo-controlled trial of two years allopurinol treatment after recent ischemic stroke or transient ischaemic attack. Visit-to-visit BPV was assessed using brachial blood pressure (BP) recordings. Short-term BPV was assessed using ambulatory BP monitoring (ABPM) performed at 4 weeks and 2 years. Brain MRI was performed at baseline and 2 years. BPV measures were compared between the allopurinol and placebo groups and with white matter hyperintensity progression.

**Results:** 409 participants were included (205 allopurinol; 204 placebo) were included in analyses of visit-to-visit BPV and there were no significant differences between groups. 196 participants were included in analyses of short-term BPV at week 4. Two measures were reduced by allopurinol: the standard deviation (SD) of systolic BP (by 1.30mmHg (95% confidence interval (CI) 0.18–2.42, p=0.023)); and the average real variability (ARV) of systolic BP (by 1.31mmHg (95% CI 0.31–2.32, p=0.011)). There were no differences in other measures at week 4 or in any measure at 2 years.

**Conclusions:** Allopurinol treatment did not affect visit-to-visit BPV in people with recent ischemic stroke or TIA. Two BPV measures were reduced at week 4 by allopurinol but not at 2 years. Allopurinol is unlikely to lead to an important reduction in BPV in people with ischemic stroke or TIA.

## Introduction

Blood pressure variability (BPV) is a strong independent predictor of cardiovascular morbidity and mortality, even after adjustment for the mean level of blood pressure (BP)^1–5^. The relationship is strongest for cerebrovascular events^1^, with high BPV being associated with both first and recurrent ischemic stroke^3, 5^, as well as worse neurologic outcome following stroke^6^. In addition, higher BPV may be associated with accelerated cognitive decline^7–9^. Elevated BPV has also been associated with the progression of imaging markers of cerebral small vessel disease, independent of mean BP^10^. Therefore, raised BPV could be a potential target for pharmacological intervention after stroke.

Hyperuricaemia is associated with an increased risk of cardiovascular morbidity and mortality^11^. A recent Mendelian randomisation analysis revealed that genetically predicted serum uric acid was associated with an increased risk of stroke, with 45% of this risk mediated through blood pressure^12^. The underlying mechanisms by which hyperuricaemia may cause hypertension include upregulation of the renin-angiotensin-aldosterone system^13^, endothelial dysfunction^14^, oxidative stress^15^, and increased arterial stiffness^16^. High serum uric acid levels have also been independently associated with increased BPV^17, 18^.

Allopurinol inhibits xanthine oxidase, resulting in reduced uric acid synthesis and serum uric acid levels. In a recent systematic review and meta-analysis, uric acid lowering treatment reduced the risk of major adverse cardiovascular events in those with prior cardiovascular disease^12^. In a recent trial, allopurinol use led to a reduction in systolic BP^19^. The effects of allopurinol on BPV remain unclear. This study aimed to investigate the effect of allopurinol on measures of BPV in people with recent ischemic stroke or transient ischemic attack (TIA) using data from the Xanthine oxidase inhibition for the Improvement of Long-term Outcomes Following Ischemic Stroke and Transient ischemic attack (XILO-FIST) trial^19^. We also aimed to assess whether there was a relationship between BPV and white matter hyperintensity (WMH) progression, brain atrophy, and cognitive function.

## Methods

XILO-FIST was a multicentre, prospective, randomised, double-blind, placebo-controlled, clinical trial of the effect of allopurinol versus placebo on cerebral WMH progression and BP in people with recent ischemic stroke or TIA. XILO-FIST was approved by the UK MHRA and NHS Research Ethics Committee (REC number 14/WS/0113) and is registered with clinicaltrials.gov (identifier: NCT02122718). All participants gave written informed consent. The trial protocol^20^ and main results^19^ have been published.

### Participant Eligibility Criteria

Participants were aged greater than 50 years and had suffered an ischemic stroke or TIA within the past 4 weeks. Full inclusion and exclusion criteria are provided in **Table S1**. Between May 2015 and December 2018, 464 participants across 22 UK sites were enrolled. Based on prior studies^4, 8^, participants were only included in the analyses for this study if they had ≥5 clinic BP readings after randomisation.

### Study Procedures

The study consisted of a 4-week run-in phase and 104-week treatment phase. Completion of the run-in phase was followed by 1:1 randomisation of participants to oral allopurinol 300mg twice daily or matching placebo for 104 weeks. Participants were reviewed at weeks 4, 13, 26, 52, 78, and 104^20^.

### BP Measurement

Brachial sphygmomanometer BP was measured at baseline and all study visits throughout the treatment phase. 24-hour ambulatory blood pressure monitoring (ABPM) was performed at baseline, and at week 4 and week 104 after randomisation unless contraindicated. Using a Spacelabs Ultralight Ambulatory Blood Pressure Monitor, readings were taken every 30 minutes during daytime (08:00–21:59) and every hour overnight (22:00–07:59). ABPM was contraindicated in participants with substantial arm weakness who would be unable to remove the device in the event of discomfort or other issues. Following the “stay-at-home” order issued by the UK government in response to the emerging COVID-19 pandemic, the XILO-FIST protocol was amended, and ABPM was no longer required at week 104.

### Brain MRI Assessment

Brain MRI was performed at baseline and week 104 using 1.5 or 3T MRI. WMHs of presumed vascular origin were defined as hyperintense lesions in the white matter on T2-FLAIR images^21^. Additional information regarding the brain MRI review process has been previously described^20^. Changes in WMH and total brain volumes were calculated as the follow-up volume minus the baseline volume.

### Cognitive Function

A Montreal Cognitive Assessment (MoCA) was performed at baseline and week 104. Change in MoCA was calculated as the visit score minus the baseline score.

### Study Outcomes

There is no current consensus on the optimal measure to estimate BPV. With substantial heterogeneity in selected measures evident between studies, the use of multiple indices is suggested^4, 6^. Visit-to-visit BPV, based on clinic BP measures, and short-term BPV, based on ABPM measures at week 4 and week 104, were assessed by 5 indices: range, standard deviation (SD), average real variability (ARV), coefficient of variation (CV), and variation independent of the mean (VIM)^22^. **Table S2** provides further details. For analysis of short-term BPV, the difference between the baseline and week 4 or 104 ABPM readings were used. Measures of short-term BPV were only calculated if the ABPM recording included ≥10 daytime and ≥5 night-time readings^23, 24^.

### Statistical analysis

Baseline characteristics were summarised. Categorical data were presented as frequency and percentages, and continuous data as mean and SD. Baseline differences were assessed using 2-sample t-tests for continuous data and chi-square tests for association for categorical data.

All analyses were conducted by intention-to-treat. 2-sample t-tests were used to determine unadjusted between group comparisons of the effect of allopurinol on visit-to-visit BPV measures. A general linear model was used to adjust for the on-treatment mean BP. The changes in short-term BPV were assessed using the same approach. We also assessed visit-to-visit BPV according to presence of hyperuricaemia at baseline (defined as serum uric acid concentration >7 mg/dL (416 μmol/L) in males and >6 mg/dL (357 μmol/L) in females^25^).

Pairwise Pearson correlation analysis was performed to examine the relationship between visit-to-visit BPV indices and changes in WMH volume, total brain volume, and MoCA.

All treatment effect estimates and correlation coefficients were reported alongside 95% confidence intervals and p-values. Tests were two-sided and a p-value of <0.05 was used for statistical significance. Analyses were conducted using SPSS 28.0 (IBM Corp., Armonk, NY) or Minitab 20.3 (Minitab Inc., State College, PA).

## Results

The trial profile is shown in **Figure 1**. 540 participants consented to participation in the trial. There were 76 withdrawals during the run-in phase and 464 participants were randomised (232 per group). Of these, 409 participants were included in the present study (205 assigned allopurinol; 204 assigned placebo – 55 participants were excluded for having <5 clinic BP measurements after randomisation). Baseline and week 4 ABPM data were available for 196 participants (96 assigned allopurinol; 100 assigned placebo). ABPM data were unavailable from 12 sites (120 participants), 43 participants did not undergo ABPM at baseline and week 4, and 50 participants had insufficient baseline and/or week 4 ABPM recordings. ABPM data were available for 115 participants at week 104 (22 recordings were missing due to the COVID-19-related protocol amendment).

**Figure 1.**
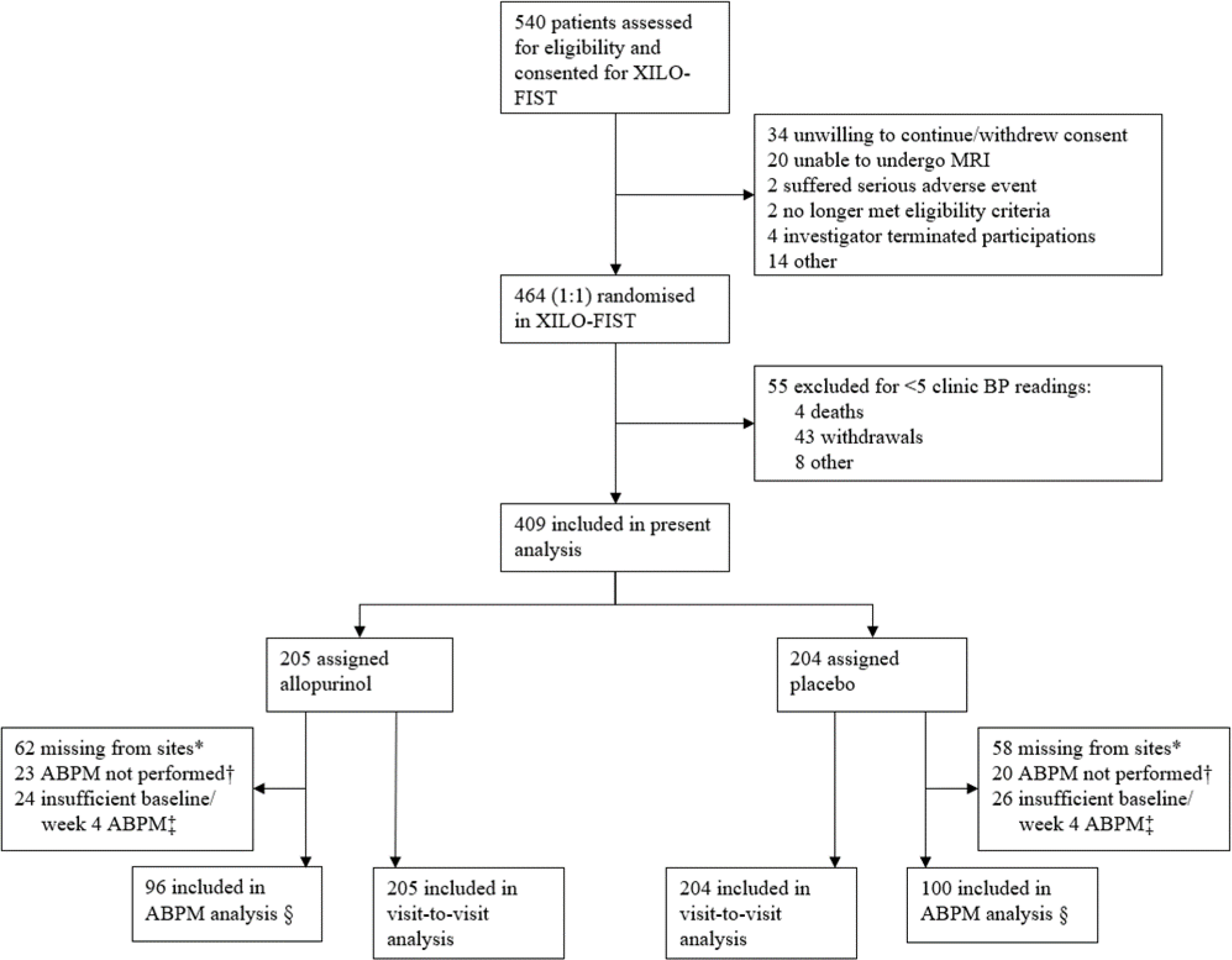
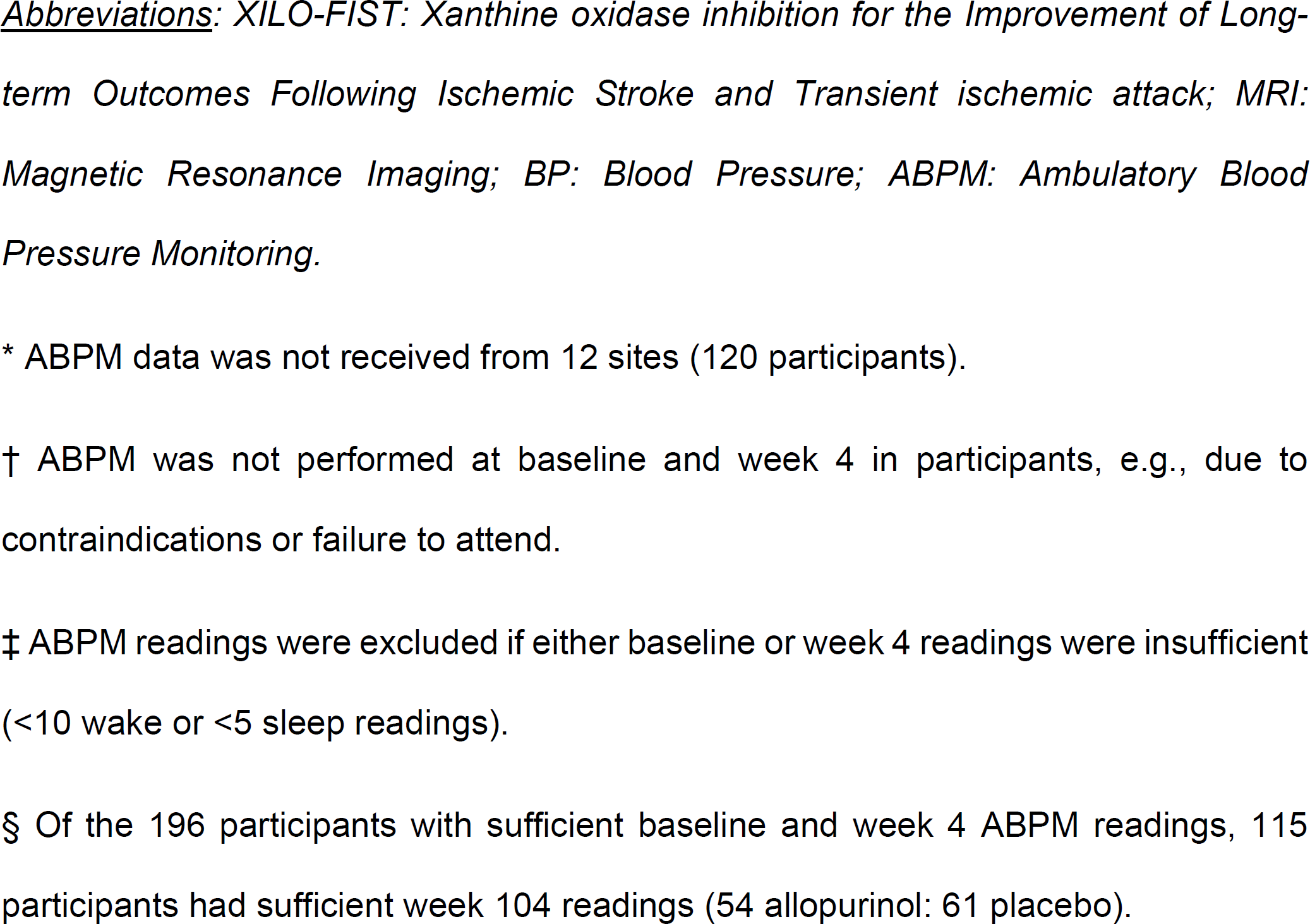
Trial Profile.

### Baseline Characteristics

Baseline demographics are shown in **Table 1**. Participants were predominantly white males (99.3% white, 69.4% male), with a mean age of 65.9 ± 8.7 years. The qualifying event was mainly ischemic stroke (93.6%; median NIHSS at enrolment = 1 (IQR: 2)). There were no significant between group differences at baseline for any measure of short-term BPV at baseline (**Table S3**). Baseline serum uric acid levels were recorded for 315 participants: 62 (19.7%) had hyperuricaemia at baseline. **Table S4** shows the baseline characteristics of participants included in this analysis and those who were excluded. Excluded participants had more severe stroke, higher BP, and greater time from index event to randomisation.

**Table 1.**
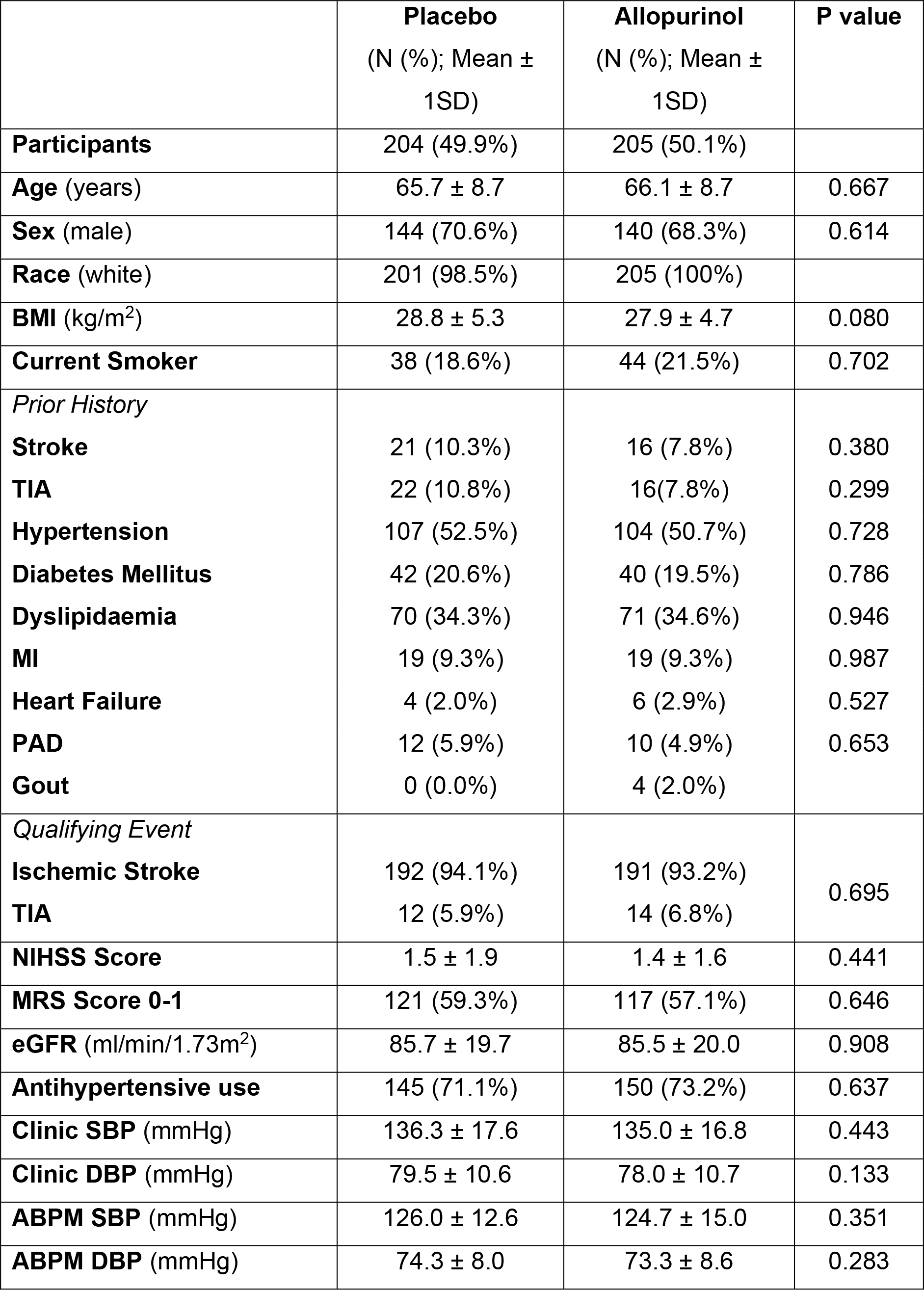
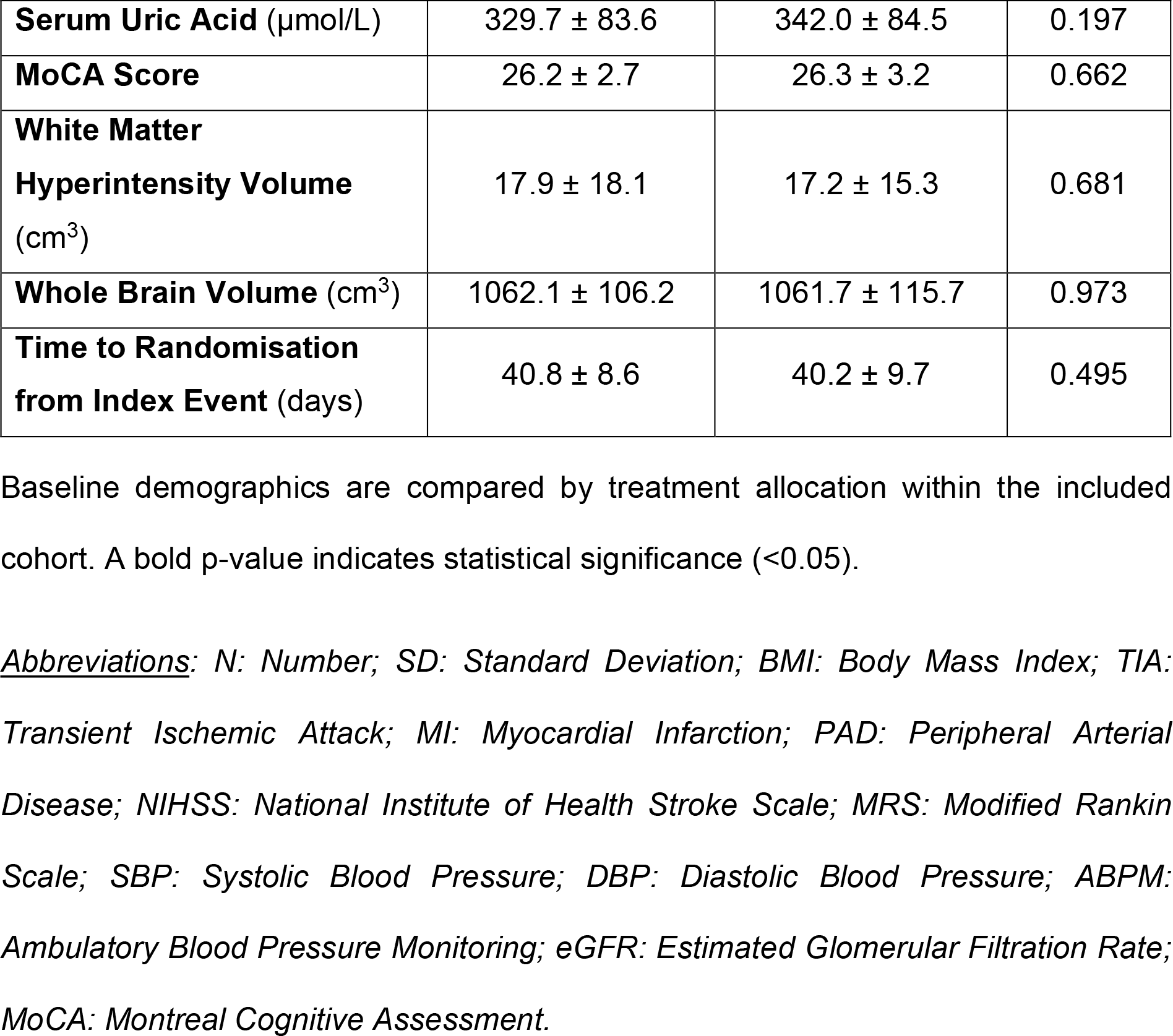
Baseline Characteristics.

|  | **Placebo**<br>(N (%); Mean ± 1SD) | **Allopurinol**<br>(N (%); Mean ± 1SD) | **P value** |
| --- | --- | --- | --- |
| **Participants** | 204 (49.9%) | 205 (50.1%) |  |
| **Age** (years) | 65.7 ± 8.7 | 66.1 ± 8.7 | 0.667 |
| **Sex** (male) | 144 (70.6%) | 140 (68.3%) | 0.614 |
| **Race** (white) | 201 (98.5%) | 205 (100%) |  |
| **BMI** (kg/m<sup>2</sup>) | 28.8 ± 5.3 | 27.9 ± 4.7 | 0.080 |
| **Current Smoker** | 38 (18.6%) | 44 (21.5%) | 0.702 |
| *Prior History* |  |  |  |
| **Stroke** | 21 (10.3%) | 16 (7.8%) | 0.380 |
| **TIA** | 22 (10.8%) | 16(7.8%) | 0.299 |
| **Hypertension** | 107 (52.5%) | 104 (50.7%) | 0.728 |
| **Diabetes Mellitus** | 42 (20.6%) | 40 (19.5%) | 0.786 |
| **Dyslipidaemia** | 70 (34.3%) | 71 (34.6%) | 0.946 |
| **MI** | 19 (9.3%) | 19 (9.3%) | 0.987 |
| **Heart Failure** | 4 (2.0%) | 6 (2.9%) | 0.527 |
| **PAD** | 12 (5.9%) | 10 (4.9%) | 0.653 |
| **Gout** | 0 (0.0%) | 4 (2.0%) |  |
| *Qualifying Event* |  |  |  |
| **Ischemic Stroke** | 192 (94.1%) | 191 (93.2%) | 0.695 |
| **TIA** | 12 (5.9%) | 14 (6.8%) |  |
| **NIHSS Score** | 1.5 ± 1.9 | 1.4 ± 1.6 | 0.441 |
| **MRS Score 0-1** | 121 (59.3%) | 117 (57.1%) | 0.646 |
| **eGFR** (ml/min/1.73m<sup>2</sup>) | 85.7 ± 19.7 | 85.5 ± 20.0 | 0.908 |
| **Antihypertensive use** | 145 (71.1%) | 150 (73.2%) | 0.637 |
| **Clinic SBP** (mmHg) | 136.3 ± 17.6 | 135.0 ± 16.8 | 0.443 |
| **Clinic DBP** (mmHg) | 79.5 ± 10.6 | 78.0 ± 10.7 | 0.133 |
| **ABPM SBP** (mmHg) | 126.0 ± 12.6 | 124.7 ± 15.0 | 0.351 |
| **ABPM DBP** (mmHg) | 74.3 ± 8.0 | 73.3 ± 8.6 | 0.283 |
| **Serum Uric Acid** (μmol/L) | 329.7 ± 83.6 | 342.0 ± 84.5 | 0.197 |
| **MoCA Score** | 26.2 ± 2.7 | 26.3 ± 3.2 | 0.662 |
| **White Matter Hyperintensity Volume** (cm<sup>3</sup>) | 17.9 ± 18.1 | 17.2 ± 15.3 | 0.681 |
| **Whole Brain Volume** (cm<sup>3</sup>) | 1062.1 ± 106.2 | 1061.7 ± 115.7 | 0.973 |
| **Time to Randomisation from Index Event** (days) | 40.8 ± 8.6 | 40.2 ± 9.7 | 0.495 |

Baseline demographics are compared by treatment allocation within the included cohort. A bold p-value indicates statistical significance (<0.05).
***Abbreviations:** N: Number; SD: Standard Deviation; BMI: Body Mass Index; TIA: Transient Ischemic Attack; MI: Myocardial Infarction; PAD: Peripheral Arterial Disease; NIHSS: National Institute of Health Stroke Scale; MRS: Modified Rankin Scale; SBP: Systolic Blood Pressure; DBP: Diastolic Blood Pressure; ABPM: Ambulatory Blood Pressure Monitoring; eGFR: Estimated Glomerular Filtration Rate; MoCA: Montreal Cognitive Assessment.*

### Visit-to-Visit BPV

Included participants had a mean of 5.9±0.3 clinic BP measurements after randomisation. No significant differences were observed between treatment groups for any measure of visit-to-visit BPV (**Table 2**). There was also no significant difference in the on-treatment mean BP between the groups. Furthermore, there were no significant differences between treatment groups for visit-to-visit BPV measures in people with and without hyperuricaemia at baseline (**Table 3**, **Table S5**).

**Table 2.**
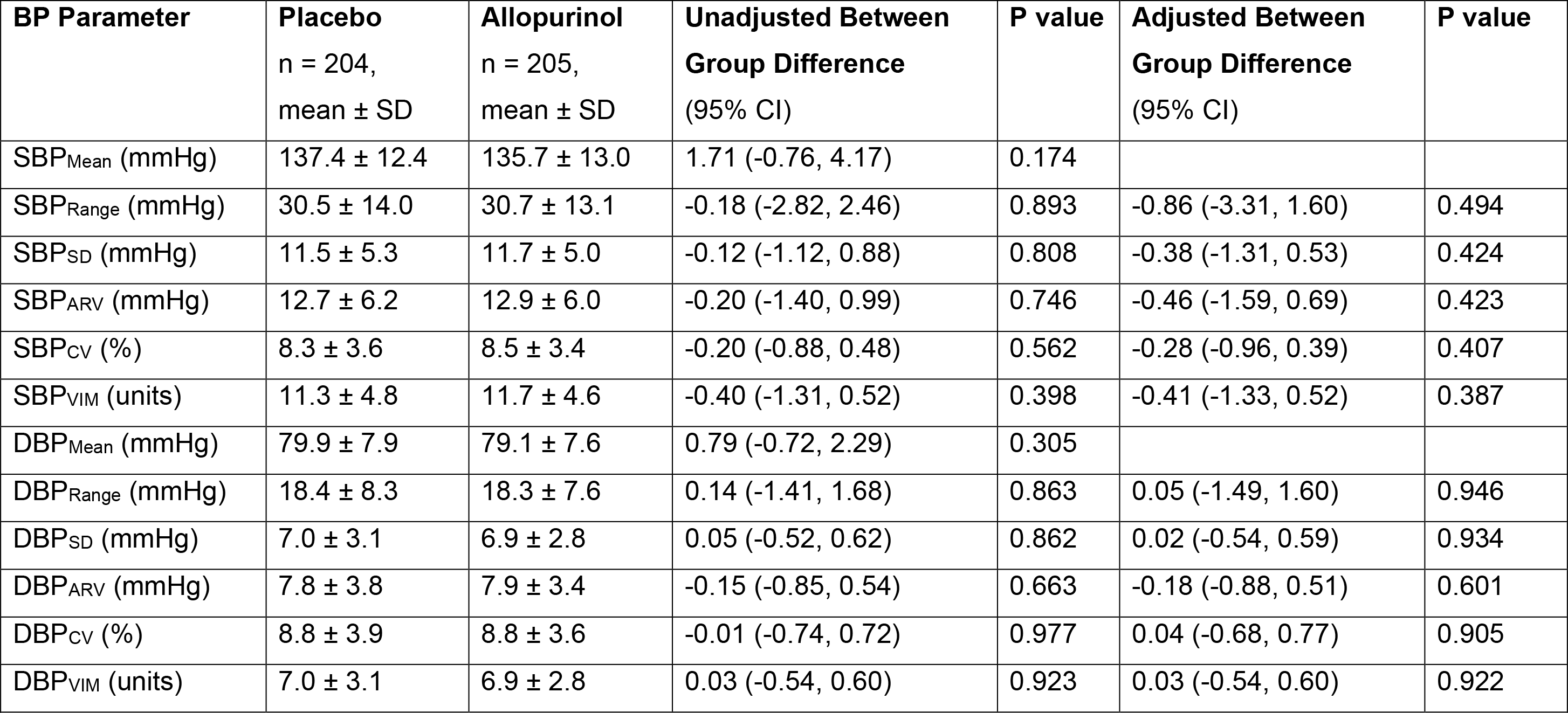
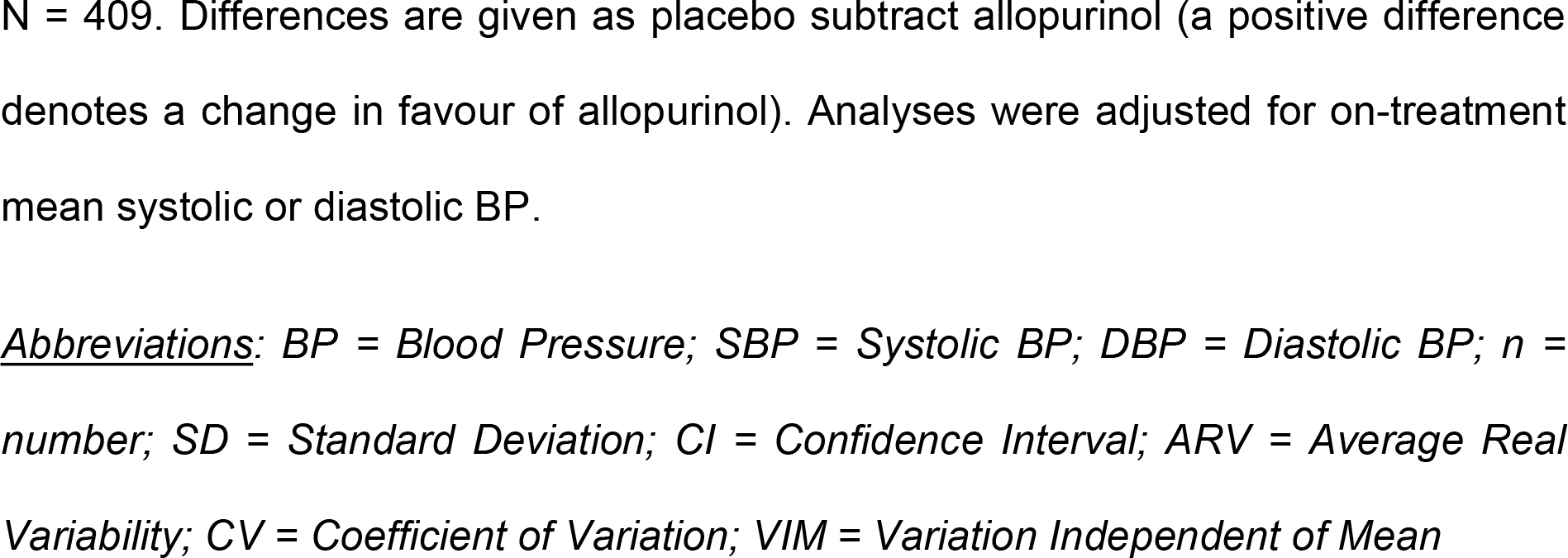
Visit-to-Visit (Clinic) Blood Pressure Variability Outcomes.

**Table 3.** Visit-to-Visit (Clinic) Blood Pressure Variability Outcomes in People with Hyperuricaemia.

| **BP Parameter** | **Placebo**<br>n = 29,<br>mean ± SD | **Allopurinol**<br>n = 33,<br>mean ± SD | **Unadjusted Between**<br>**Group Difference**<br>(95% CI) | **P**<br>**value** |
| --- | --- | --- | --- | --- |
| SBP<sub>Range</sub> (mmHg) | 32.3 ± 12.1 | 35.1 ± 14.2 | -2.86 (-9.52, 3.80) | 0.394 |
| SBP<sub>SD</sub> (mmHg) | 12.4 ± 4.8 | 13.3 ± 5.1 | -0.98 (-3.48, 1.53) | 0.440 |
| SBP<sub>ARV</sub> (mmHg) | 13.7 ± 5.3 | 14.8 ± 6.1 | -1.07 (-3.96, 1.83) | 0.464 |
| SBP<sub>CV</sub> (%) | 9.2 ± 3.5 | 9.5 ± 3.5 | -0.35 (-2.12, 1.41) | 0.691 |
| SBP<sub>VIM</sub> (units) | 12.7 ± 4.8 | 12.9 ± 4.7 | -0.12 (-2.54, 2.29) | 0.919 |
| DBP<sub>Range</sub> (mmHg) | 17.7 ± 8.7 | 19.7 ± 7.1 | -2.02 (-6.10, 2.05) | 0.324 |
| DBP<sub>SD</sub> (mmHg) | 6.7 ± 3.3 | 7.4 ± 2.5 | -0.72 (-2.24, 0.80) | 0.343 |
| DBP<sub>ARV</sub> (mmHg) | 7.1 ± 3.4 | 8.5 ± 2.9 | -1.38 (-3.00, 0.24) | 0.093 |
| DBP<sub>CV</sub> (%) | 8.8 ± 4.6 | 9.3 ± 3.2 | -0.51 (-2.55, 1.53) | 0.618 |
| DBP<sub>VIM</sub> (units) | 6.8 ± 3.4 | 7.4 ± 2.5 | -0.61 (-2.15, 0.94) | 0.434 |

N = 62. Differences are given as placebo subtract allopurinol (a positive difference denotes a change in favour of allopurinol).
***Abbreviations:** BP = Blood Pressure; SBP = Systolic BP; DBP = Diastolic BP; n = number; SD = Standard Deviation; CI = Confidence Interval; ARV = Average Real Variability; CV = Coefficient of Variation; VIM = Variation Independent of Mean*

### Short-term (ABPM) BPV Outcomes

Participants had a mean of 29.6±6.4 (21.2±6.1 daytime and 8.4±1.8 night-time) BP readings taken during week 4 ABPM. The changes in measures of short-term (ABPM) BPV from baseline to week 4 are shown in **Table 4**. At week 4, there were no significant differences in the change of mean BP or diastolic BPV between treatment groups. There were no differences in the change in the range, CV, and VIM of systolic BP (SBP) between groups. The change in SD of SBP was lower with allopurinol compared to placebo, with a between group difference of 1.30mmHg (95% CI 0.18– 2.42, p=0.023, p=0.051 after adjustment for change in mean SBP). The change in ARV of SBP was also lower with allopurinol, with a difference between groups of 1.31mmHg (95% CI 0.31–2.32, p=0.011, p=0.028 after adjustment for change in mean SBP (p=0.028).

**Table 4.**
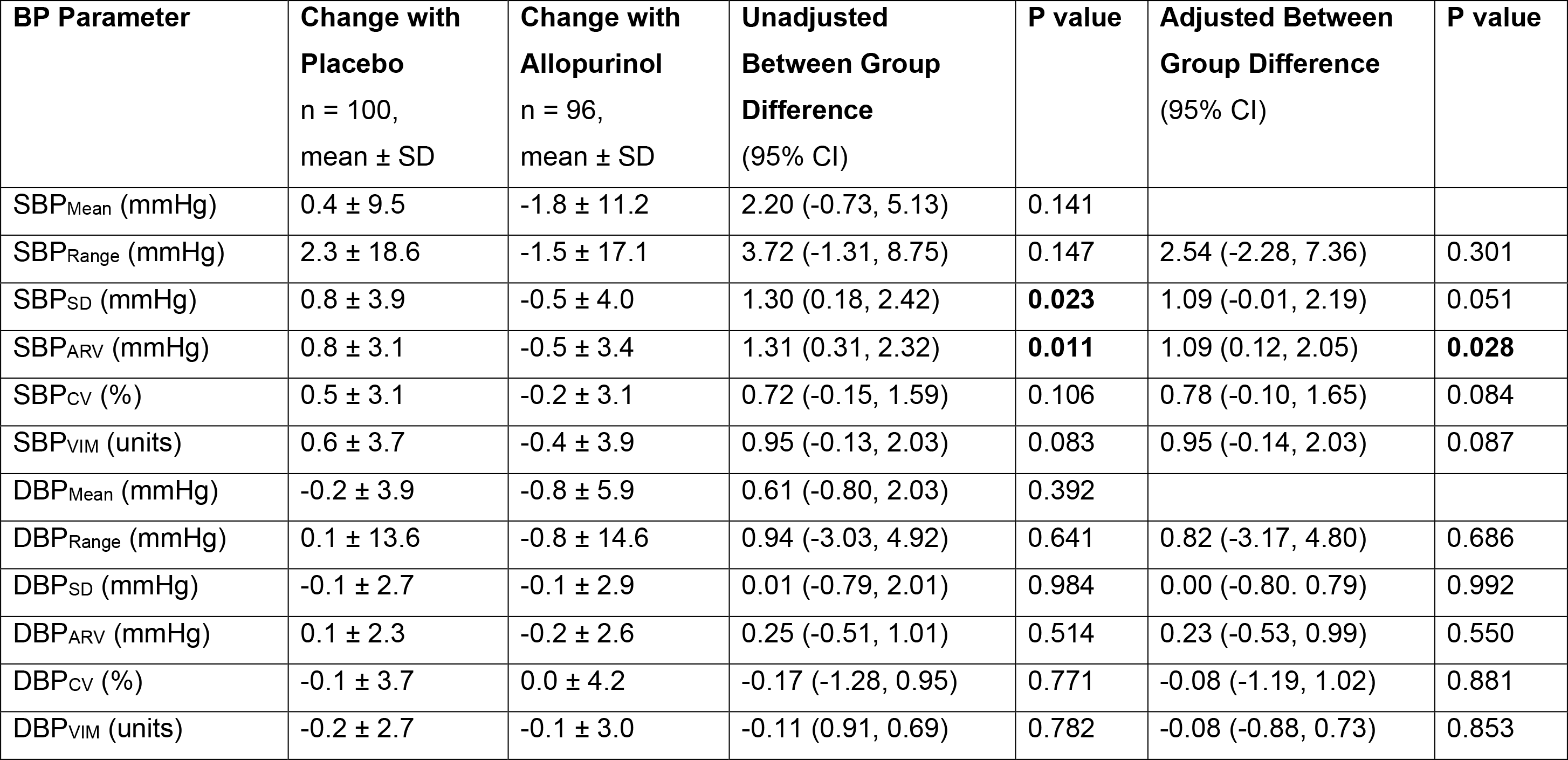
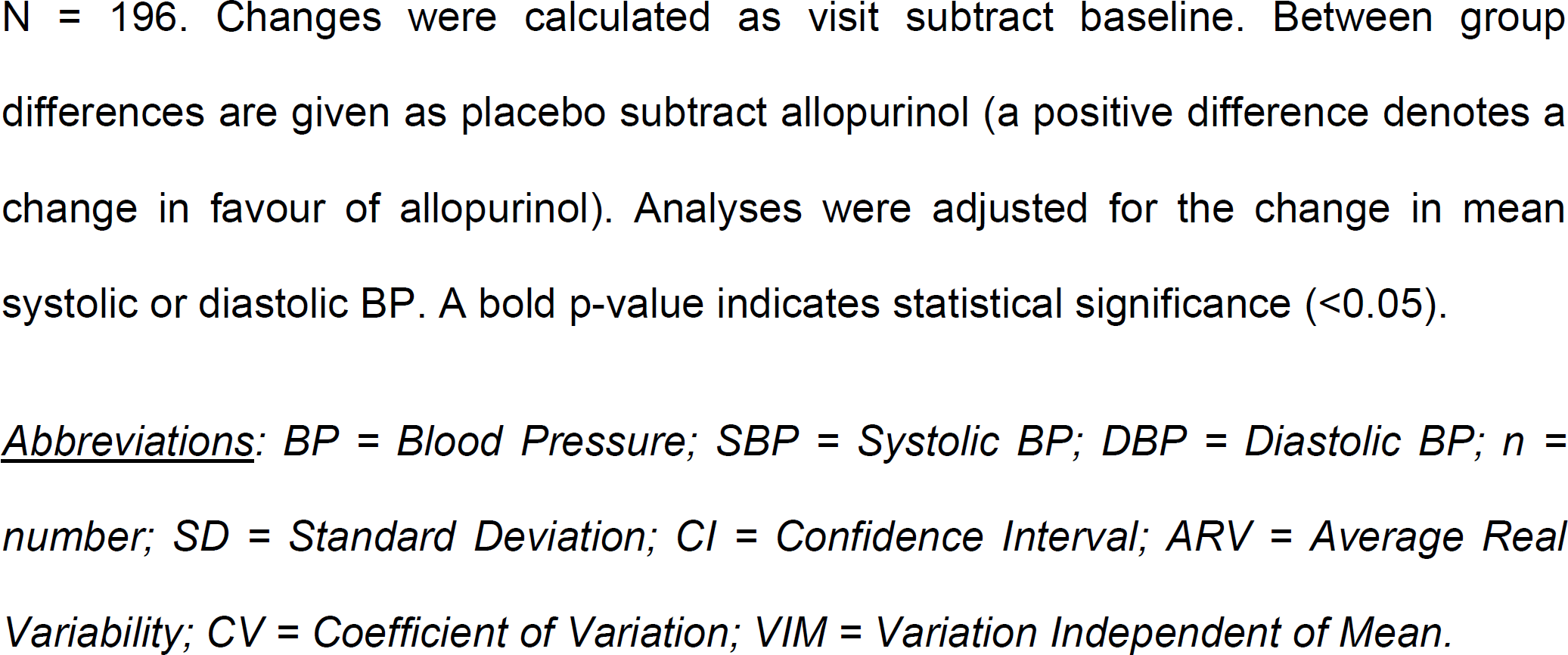
Change in Short-term (ABPM) Blood Pressure Variability from Baseline to Week 4.

At week 104, participants had a mean of 27.7±5.2 (19.7±4.8 daytime and 8.2±1.5 night-time) BP readings taken during ABPM. There were no significant differences between treatment groups for the change in any measure of systolic or diastolic short-term BPV at week 104 (**Table 5**).

**Table 5.**
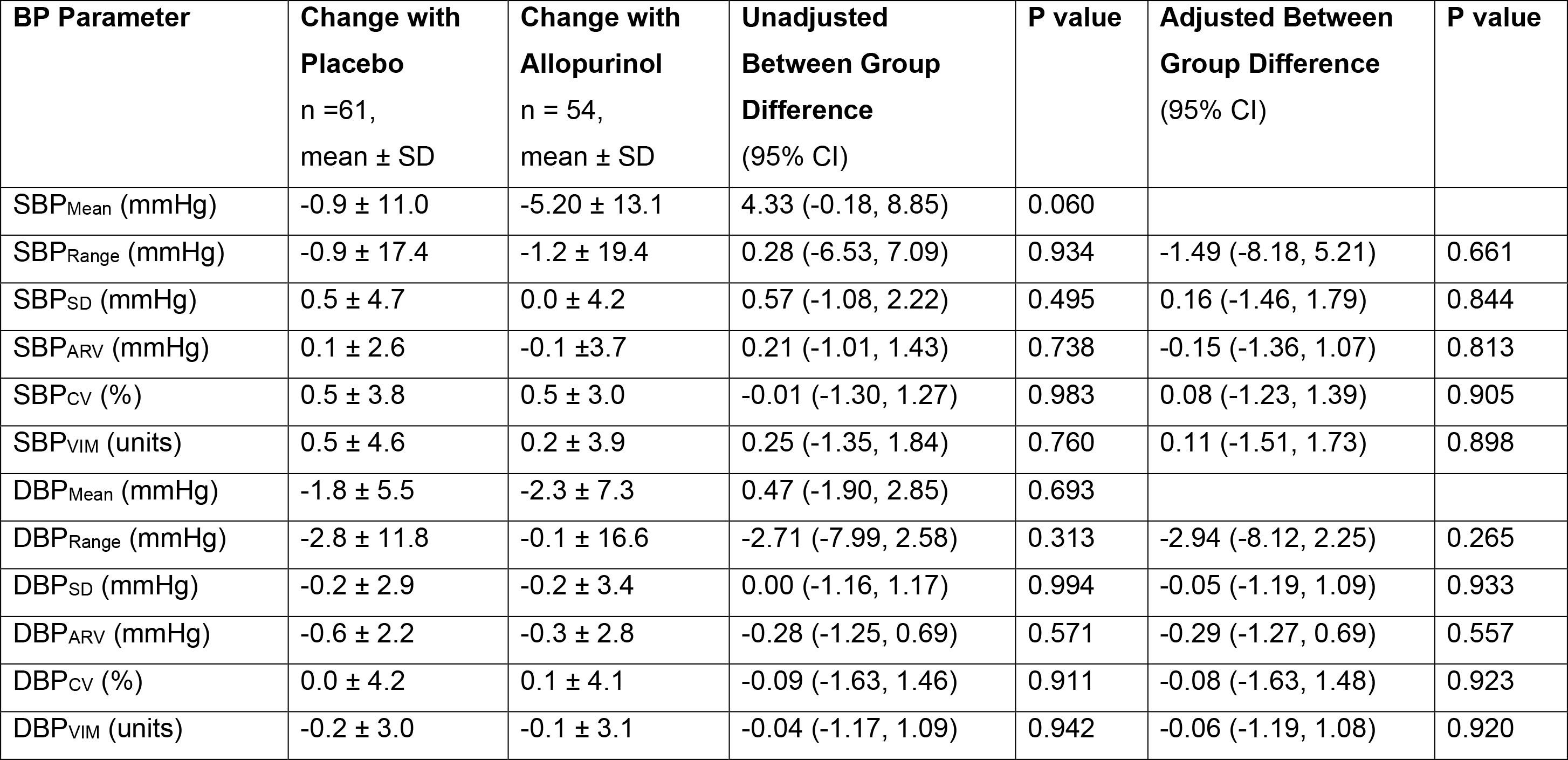
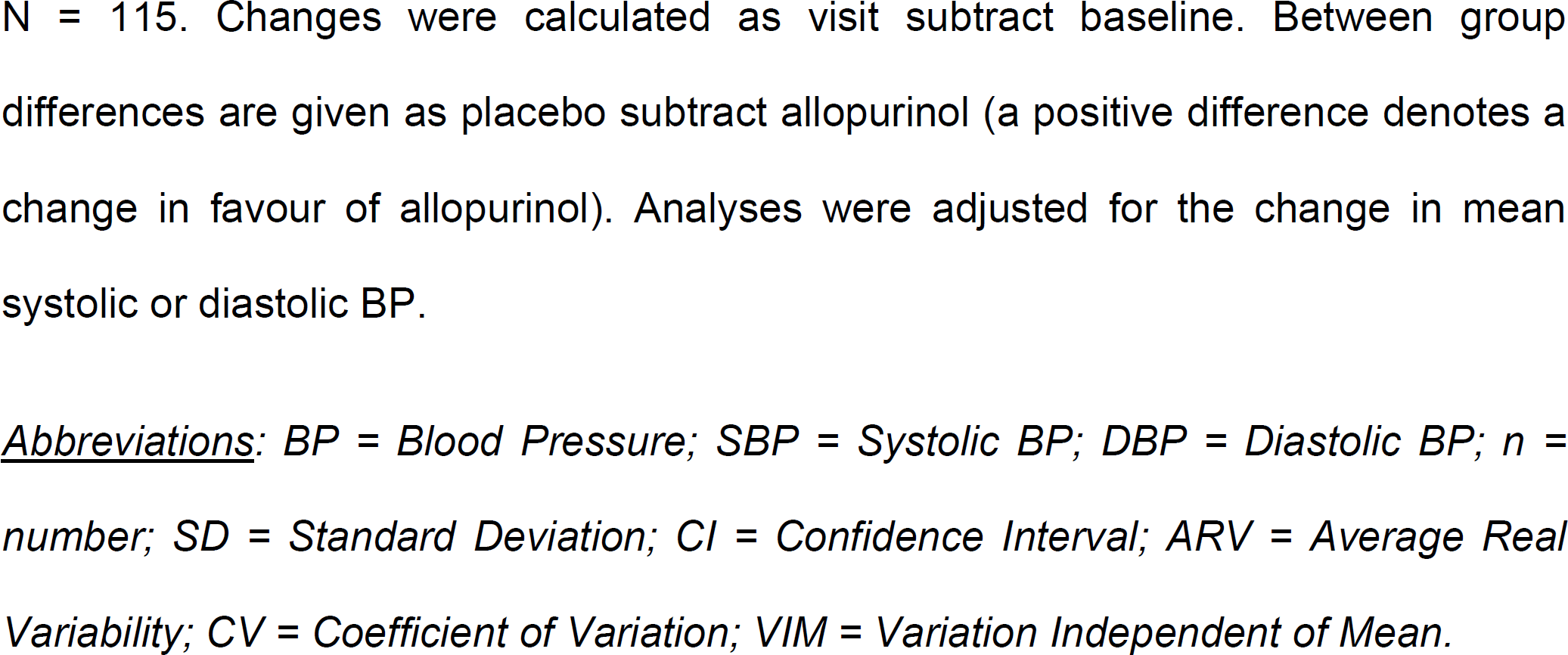
Change in Short. -term (ABPM) Blood Pressure Variability from Baseline to Week

### BPV, White Matter Hyperintensity Progression, Brain Atrophy and Cognition

A total of 367 participants (181 assigned allopurinol; 186 assigned placebo) had visit-to-visit BPV data and WMH volume assessments performed at baseline and week 104. The median change in WMH volume was a 1.02cm^3^ increase (IQR: 3.66). Visit-to-visit BPV was not significantly associated with change in WMH volume across any measure of BPV (**Table S6**). Brain volume measurements could be performed on 359 participants (179 assigned allopurinol; 180 assigned placebo). The median change in total brain volume was a 17.29cm^3^ decrease (IQR: 28.13). Visit-to-visit BPV was not significantly associated with change in brain volume across any measure of BPV (**Table S7**).

A total of 368 participants (180 assigned allopurinol; 188 assigned placebo) had visit-to-visit BPV data and MoCA performed at baseline and week 104. 34.0% experienced a decline in their MoCA score from baseline. Visit-to-visit BPV was not significantly associated with change in MoCA across any measure of BPV (**Table S8**).

## Discussion

In this exploratory analysis of the XILO-FIST trial, we investigated the effect of allopurinol on BPV in people with recent ischemic stroke or TIA. The main findings were as follows: (i) 2-year treatment with allopurinol did not significantly affect any measure of clinic visit-to-visit BPV compared to placebo; (ii) allopurinol treatment reduced the SD and ARV of SBP from baseline to follow-up at week 4, and the change in ARV remained significant after adjusting for the change in mean SBP; (iii) there were no significant differences for the change in any other measure of systolic or diastolic short-term BPV from baseline to week 4 between treatment groups; and (iv) there were no significant differences between groups for the change in short-term BPV from baseline to study end at week 104. Furthermore, we investigated the relationship between BPV and WMH progression, brain atrophy, and cognitive function. No visit-to-visit BPV measure was associated with progression of WMH volume, brain atrophy, or cognitive decline in this study.

Despite previously reported associations between hyperuricaemia and BPV^17, 18^, we conclude that allopurinol does not affect clinic visit-to-visit BPV in people with recent stroke or TIA. Where an association has been seen between hyperuricaemia and BPV, the population studied has typically been younger and healthier than those in our study^17, 18^. Although BP was lowered by allopurinol in the XILO-FIST trial, prior clinical trials suggest greater effect of uric acid lowering therapy on BP in younger, pre- or newly-hypertensive populations^26, 27^, compared with older adults, especially when people are already receiving anti-hypertensive treatment^28^. This is in-keeping with experimental data which postulates that hyperuricaemia-induced hypertension is initially reversible with uric acid lowering agents^13^, but with time the hypertension becomes salt-sensitive and resistant to uric acid lowering therapy^29^. The relationship between uric acid and BPV may follow a similar pattern.

Compared to placebo, allopurinol treatment reduced short-term BPV measured by the SD and ARV of SBP from baseline to week 4 by 1.3mmHg. The absolute magnitude of this difference is greater than the differences observed in the X-CELLENT study^30^, where amlodipine reduced SBP SD by 1.0mmHg and ARV by 0.6mmHg versus placebo. However, after adjusting for the change in mean SBP, the effect of allopurinol on the change in SD of SBP was not significant. In contrast, the change in ARV remained significant. This may be explained by the fact that ARV is more sensitive to the sequential order of BP measurements and less sensitive to the influence of low sampling frequency or day-to-night differences (e.g., the nocturnal BP fall which can confound SD)^2, 22, 31^. In addition, ARV has been shown to offer greater prognostic value to ABPM than SD^2^ and closer correlations have been identified between markers of arterial stiffness and ARV than SD^2^. This may perhaps explain the greater effect of allopurinol on the ARV of short-term BPV compared to the SD. Furthermore, there were no differences in any measure at week 104. The difference in results at week 104 could be due to type 2 error. It is also possible that the differences seen at week 4 could be due to type 1 error, because of the multiple comparisons made.

We investigated the relationship between BPV, WMH progression, brain atrophy, and cognitive function. Visit-to-visit BPV was not associated with change in WMH volume in our study. This contrasts with the findings of a recent systematic review and meta-analysis which linked BPV with WMH burden^10^. However, this meta-analysis did not include studies with high numbers of people with stroke. Consistent with our findings, Liu et al.^32^ found that no visit-to-visit BPV parameter predicted WMH progression in people with a history of ischemic stroke. Likewise, despite previously documented links between elevated BPV, brain atrophy and cognitive decline^7–9, 33, 34^, we found no relationship between measures of visit-to-visit BPV and change in brain volume or cognitive decline in this post-stroke population.

As an exploratory analysis, a key strength of our study is the multicentre, randomised, double-blind, placebo-controlled design of XILO-FIST. However, our study has important limitations. First, ABPM data was not received for 120 participants. We required access to the raw data for this analysis and this was not possible due to COVID-19 restrictions which prevented us from travelling to each individual health board to extract the data. This cannot be done now as the study is complete. Second, participants were predominantly white males which may limit the generalisability of our findings. Third, the issue of multiplicity must be considered. Since this analysis was hypothesis generating, we did not adjust for multiple testing.

### Perspectives

Allopurinol treatment did not affect visit-to-visit BPV in people with recent ischemic stroke or TIA. Allopurinol may reduce short-term BPV measured by the SD and ARV of systolic BPV over a short-term time horizon, although it is unlikely to lead to a sustained clinically important change in BPV.

### Novelty and Relevance

What Is New?

- We did not find evidence of an effect of high dose allopurinol on BPV in people with previous stroke or TIA.

What Is Relevant?

- People with stroke and TIA are at high risk of recurrent events.
- High serum uric acid levels are associated with increased risk, which is partly mediated via a higher blood pressure. It may also be associated with BPV.
- Allopurinol has been shown to lower blood pressure and might have an effect on BPV.

Clinical/Pathophysiological Implications?

- Our data provide further evidence that urate lowering therapy with allopurinol is unlikely to reduce recurrence risk in people with ischaemic stroke or TIA.

## Data Availability

The datasets analysed that support the findings of the present study are available from the corresponding author upon reasonable request.

## Acknowledgments

We would like to thank all the XILO-FIST investigators, study staff and participants.

## Disclosures

AS holds a patent for the use of xanthine oxidase inhibition for the treatment of angina pectoris.

DD received payment for image analysis in this study.

The other authors declare they have no competing interests.

## Supplementary Material

Tables S1-8

## Non-standard Abbreviations and Acronyms

ABPM: Ambulatory blood pressure monitoring
ARV: Average real variability
BMI: Body mass index
BP: Blood pressure
BPV: Blood pressure variability
CI: Confidence interval
CV: Coefficient of variation
DBP: Diastolic blood pressure
IQR: Interquartile Range
MoCA: Montreal cognitive assessment
MRI: Magnetic resonance imaging
NIHSS: National institute of health stroke scale
SBP: Systolic blood pressure
SD: Standard deviation
TIA: Transient ischemic attack
VIM: Variation independent of the mean
WMH: White matter hyperintensity
XILO-FIST: Xanthine oxidase inhibition for the improvement of long-term outcomes following ischemic stroke and transient ischemic attack

